# Relative contributions of vaccination and previous infection to population-level SARS-CoV-2 immunity over time: a simulation modelling study

**DOI:** 10.1101/2022.08.18.22278963

**Authors:** Joshua Szanyi, Tim Wilson, Hassan Andrabi, Tony Blakely

## Abstract

Population-level immunity to SARS-CoV-2 directly impacts the incidence of COVID-19 morbidity and mortality. Understanding how this immunity is likely to change over time in the context of future vaccination schedules and emerging SARS-CoV-2 variants is critical to inform pandemic policy. This study simulates population-level COVID-19 immunity (including relative contributions of vaccination and previous infection) in Victoria, Australia over 18 months using an agent-based model and logistic regression equations that predict immunity and waning following vaccination and/or infection. Previous infection was found to drive most immunity against infection even with ongoing regular vaccination, however a greater proportion of overall immunity against mortality was accounted for by vaccination. Although previous infection appears to be driving a substantial component of population-level COVID-19 immunity currently, improved vaccines providing longer lasting (and better sterilizing) immunity are likely to be a critical component of the future pandemic response given the risks associated with SARS-CoV-2 infection.

## Introduction

The emergence of immune-evasive variants, waning immunity, and relatively poor sterilizing immunity afforded by vaccination mean that currently available vaccines have had less of an impact on the long-term course of the COVID-19 pandemic than hoped. Protection against symptomatic Omicron BA.1 infection following a third dose of an mRNA vaccine, for example, peaks at approximately 60% – 70% and wanes rapidly thereafter; ^1^ protection against more severe clinical outcomes is higher but still decreases over time. ^2^ The Omicron sub-variants BA.4/BA.5 possess even greater immune evasion capacity than earlier sub-variants, ^3^ highlighting the challenge of responding to an ever-evolving virus under strong selection pressure to evade pre-existing immune responses in the population. ^4^

Where the primary infection and subsequent re-exposure are due to the same SARS-CoV-2 variant, the level of immunity that develops following infection appears to be high, particularly among younger people. ^5^ It also appears to wane at a slower rate than vaccine-derived immunity. ^5^ Hybrid immunity (vaccination augmented by natural infection) provides additional protection above either sensitizing event alone.^6^

COVID-19-related morbidity and mortality is influenced by the level of immunity in a population at any given point in time. Accordingly, it is important to understand total population-level immunity, the relative contributions of vaccination and previous infection to this immunity, and how this is likely to change to inform ongoing pandemic policy such as vaccine scheduling. This is a challenge, however, given the current dynamic and mixed picture of vaccination and infection-derived immunity in many jurisdictions in addition to imperfect knowledge regarding expected protection against future variants.

We previously developed a logistic regression model of vaccine effectiveness (VE) that provides estimates of protection and waning against symptomatic infection and hospitalisation with the Omicron BA.1 variant. ^7^ We subsequently extended this equation, incorporating it into an agent-based model (ABM) to assess optimal pandemic policy in the context of ongoing SARS-CoV-2 evolution over an 18-month period in Victoria, Australia. ^8^

The current study aims to use this ABM to simulate total population-level immune protection against SARS-CoV-2 in Victoria over the same period, and the relative contributions of vaccination and previous infection to this protection given four illustrative COVID-19 vaccination schedules and eight potential SARS-CoV-2 variant scenarios.

## Methods

VE estimates were based on a previously published logistic regression model that estimated waning protection against symptomatic infection and hospitalisation from the Omicron BA.1 variant. ^7^ We extended this initial equation to account for age using data from a study reporting age-stratified VE against the Delta variant. ^9^ We then added the outcomes of death and any infection (using UK Health Security Agency data). ^2^ Finally, we developed a similar model to represent protection following previous infection (that is, protection against reinfection with the same SARS-CoV-2 variant upon re-exposure) using the same relative differences in protection across clinical outcomes as for VE and waning set halfway between nil waning and that for VE against hospitalisation. ^7^ Hybrid immunity was calculated as 1 – (1 – VE) × (1 – protection from natural infection) based on evidence suggesting that vaccine- and infection-derived immunity act independently to provide an overall level of protection. ^6^ For further details, please see the Supplementary Materials of ^8^.

These immunity estimates were then assigned to agents in an ABM that was developed to assess optimal pandemic policy in the state of Victoria, Australia (6.6 million people), in the 18 months from 1 April 2022.^8^ For the current study, four vaccine schedules were included – nil further vaccination beyond 1 April 2022; three 6-monthly boosters of current-generation mRNA vaccines (with rollout from July 2022, January 2023 and July 2023); two doses of an illustrative next-generation Omicron-targeted vaccine (with rollout from October 2022 and April 2023); and two doses of an Omicron-targeted vaccine with its waning rate set at 25% of that for current vaccines (Table 1). Relative VE of the Omicron-targeted vaccine against the SARS-CoV-2 variants modelled in the current study was set using an odds ratio of 2 applied to current-generation VE estimates (Supplementary Table 3; by way of illustration, a doubling of VE on the OR scale means a VE of 90% becomes 95%, and 40% become 57%).

**Table 1:** Modelled vaccination schedules

| Schedule | Apr-Jun<br>2022 | Jul-Sep<br>2022 | Oct-Dec<br>2022 | Jan-Mar<br>2023 | Apr-Jun<br>2023 | Jul-Sep<br>2023 |
| --- | --- | --- | --- | --- | --- | --- |
| a) Nil further |  |  |  |  |  |  |
| b) 3 current generation (CG) |  | CG |  | CG |  | CG |
| c) 2 Omicron-targeted (OT) |  |  | OT |  | OT |  |
| d) 2 OT (with 25% waning) |  |  | OT |  | OT |  |

*Applies to ≥20-year-olds with triple dose coverage and <20-year-olds with double dose coverage on 1 April 2022. At each wave of vaccination, it is assumed that 80% of people (+/-10% SD, beta distribution) vaccinated in the last wave take up the new offering. For example, 80% of ≥20-year-olds who start the model on 1 April 2022 already triple-vaccinated receive the next dose of the scheduled vaccine. ≥20-year-olds who had not received their third dose by 1 April 2022 do not receive any further vaccines. Vaccine rollout is even over three months with no prioritization by age.*

At each round of vaccination, 80% of agents vaccinated in the previous round were vaccinated with the new vaccine. Vaccine schedules were modelled in the context of eight potential future SARS-CoV-2 variants, all with incursion into Victoria 91 days into the model run, with two potential levels of innate transmissibility (R0 11 or 14), immune escape capacity and virulence (low, approximating the Omicron variant, or high) ^8^. Immune escape possibilities for each SARS-CoV-2 variant are shown in Supplementary Table 3 and Supplementary Table 4. The virus circulating at model outset, prior to the emergence of a new variant, approximated Omicron BA.2.

In the ABM, public health and social measures (including physical distancing and mask wearing) (de)escalated through five stages based on expected health service pressure (Supplementary Table 1 and Supplementary Table 2). Over the 18-month period modelled, weekly SARS-CoV-2 infections and deaths due to COVID-19 were averaged across 100 separate draws of model input parameters, each run four times to dampen stochastic variation. To illustrate variability across the 100 model runs, 5^th^ and 95^th^ percentiles of weekly infections and deaths are also presented in the results; the purpose of this study was not to precisely predict epidemic dynamics in the short-term but explore the cumulative impact of various pandemic policies given a highly uncertain future.

Population-level immunity was quantified as the average percentage reduction (compared to nil vaccination and nil previous infection) of the probability of the event of interest (being infected, death if infected) across all agents. This average immunity was calculated from vaccines alone, previous infection alone, and the total from both vaccination and previous infection (including hybrid immunity).

## Results

Figure 1 shows total population immunity over 18 months for all modelled viral variants, in the context of 6-monthly boosters with a current generation mRNA vaccine. Total immunity varied depending on the variant, particularly for protection against infection. Lower levels of immunity against infection were seen with more highly virulent variants; higher levels of total immunity were seen with more transmissible variants possessing lower virulence.

**Figure 1:**
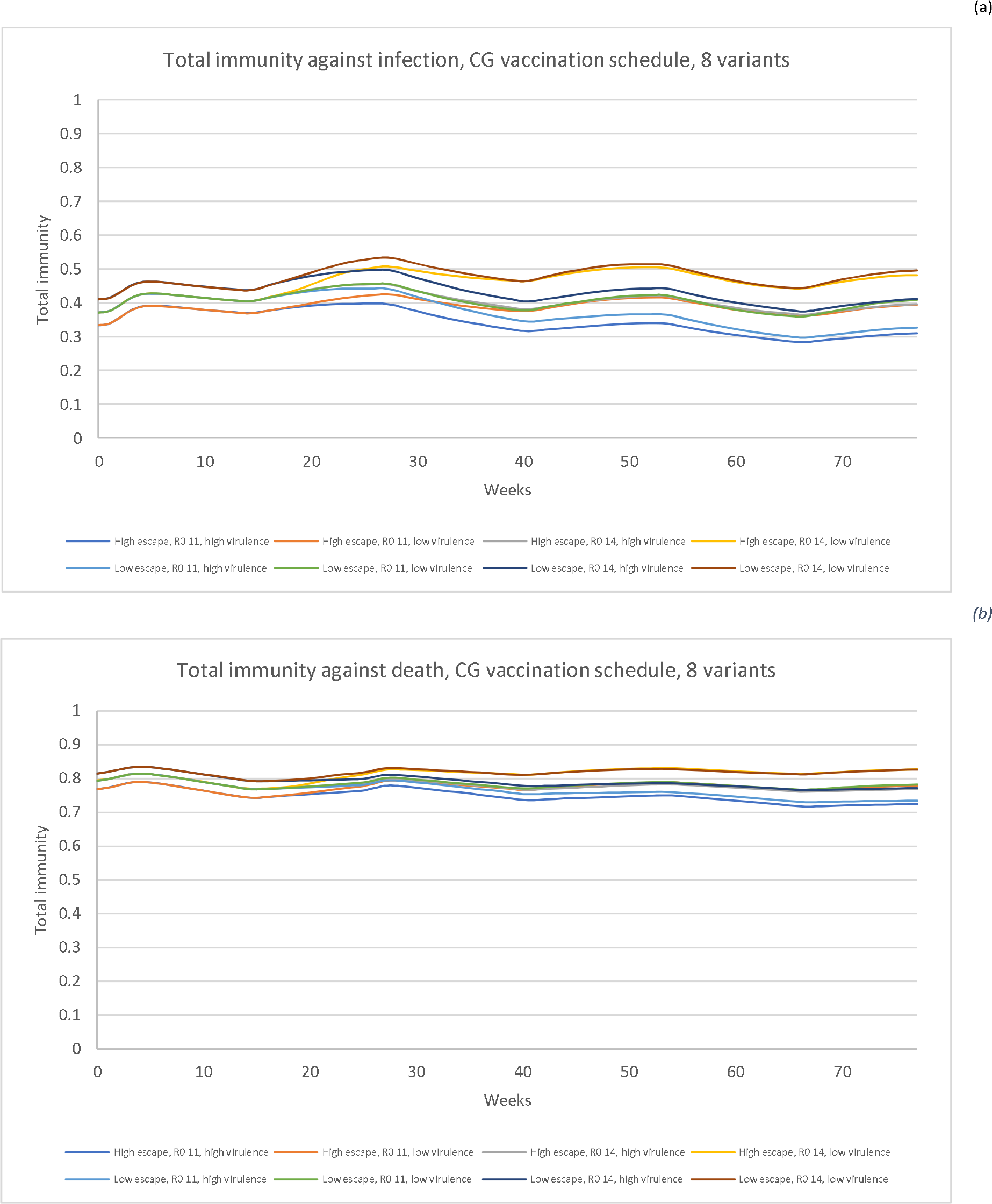
Simulated total immunity against (a) any infection and (b) death for 6-monthly boosters with a current generation vaccine across eight modelled SARS-CoV-2 variants*. * Variant incursion at 13 weeks. See Table 1 for vaccination timing.

Irrespective of the vaccination scenario, population-level immunity reached a relatively steady state, but with more peaks and troughs in scenarios with ongoing vaccination. This is evident in Figure 2 and Figure 3, which show simulated total immunity, vaccine-derived immunity, and immunity from previous infection against any infection (symptomatic and asymptomatic infection) and death for one future

**Figure 2:**
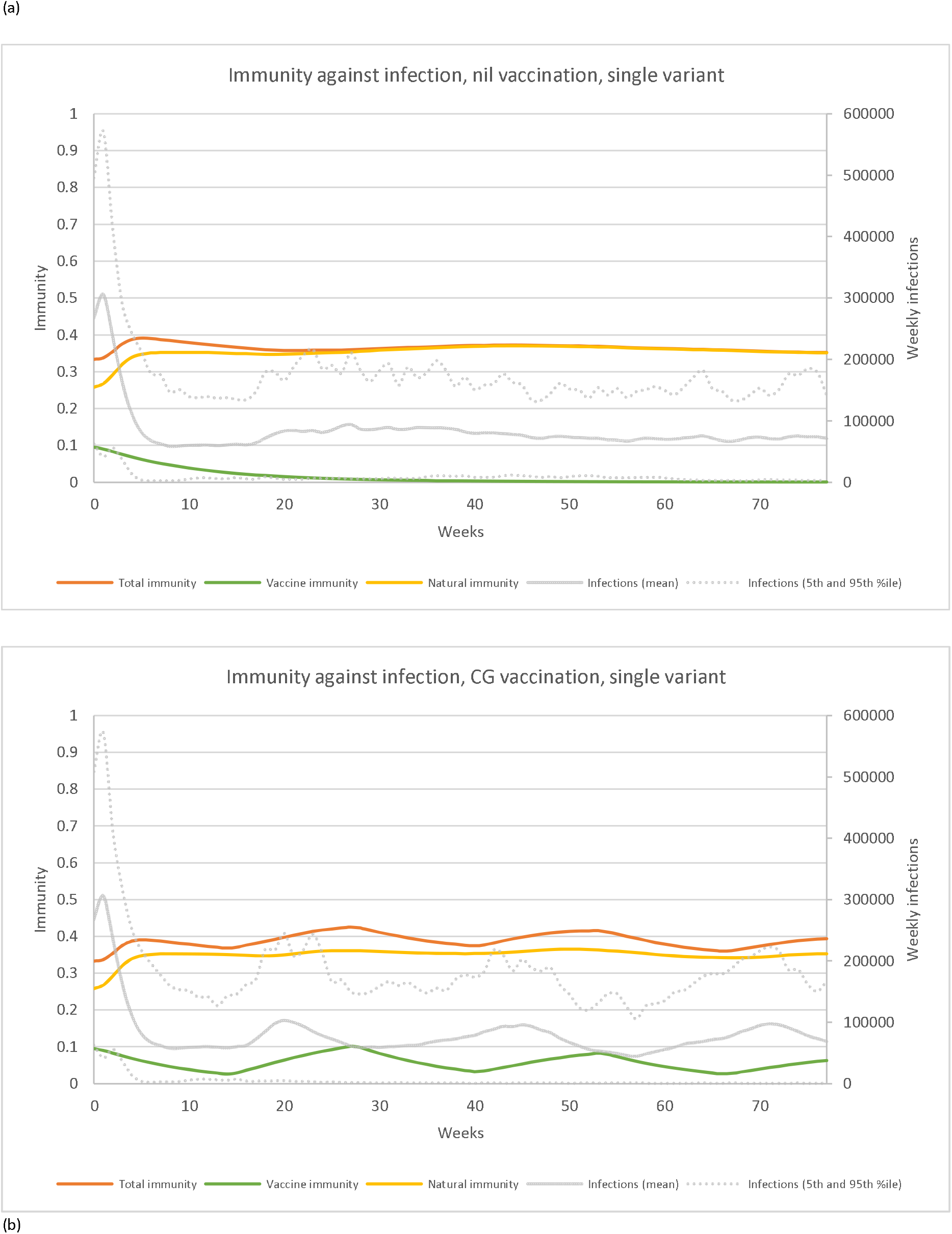
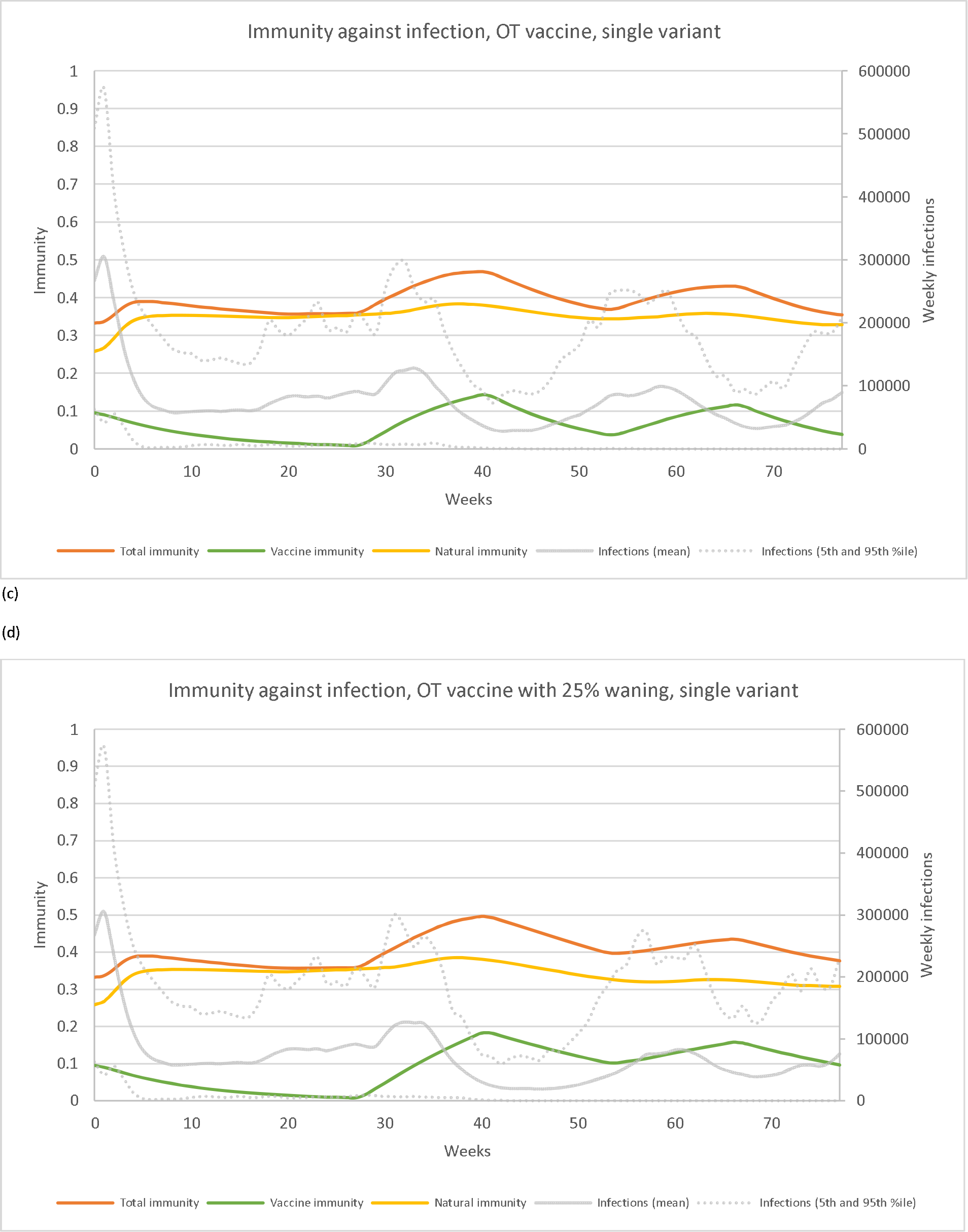
Simulated total immunity, vaccine-derived immunity, and immunity from previous infection against <u>any infection</u> over an 18-month period in Victoria, Australia for one future SARS-CoV-2 variant* and (a) nil further vaccination, (b) 6-monthly boosters with a current generation vaccine, (c) two doses of an Omicron-targeted next-generation vaccine with the same waning as current vaccines, and (d) two doses of an Omicron-targeted vaccine with 75% less waning than current vaccines. * variant with R_0_ 11, antigenically Omicron-like, virulence approximating the Omicron variant, emerging in July 2022 (13 weeks), with high immune escape capacity (halving vaccine effectiveness and/or natural protection on the odds ratio scale). See Table 1 for vaccination timing. CG: current generation; OT: Omicron-targeted.

**Figure 3:**
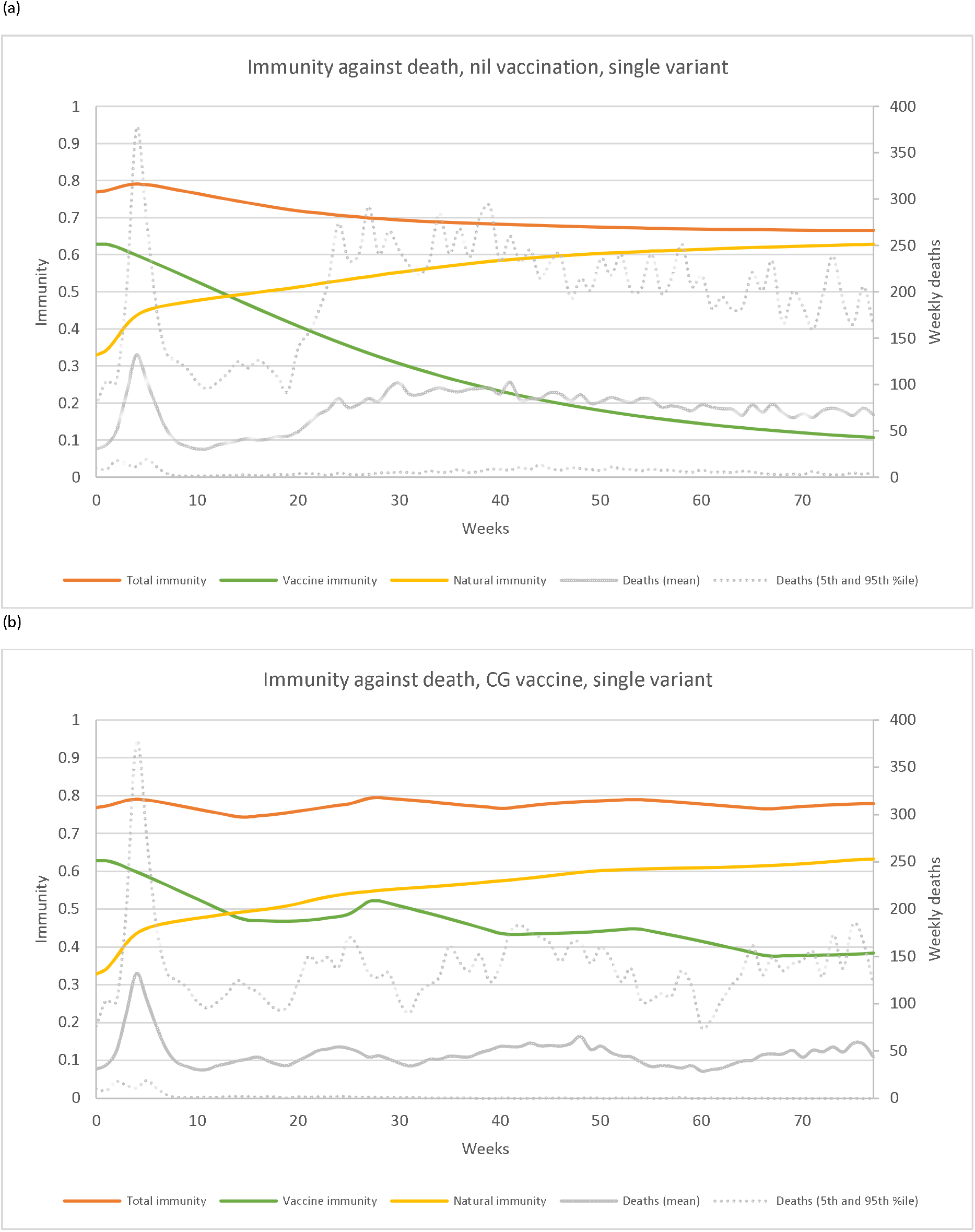
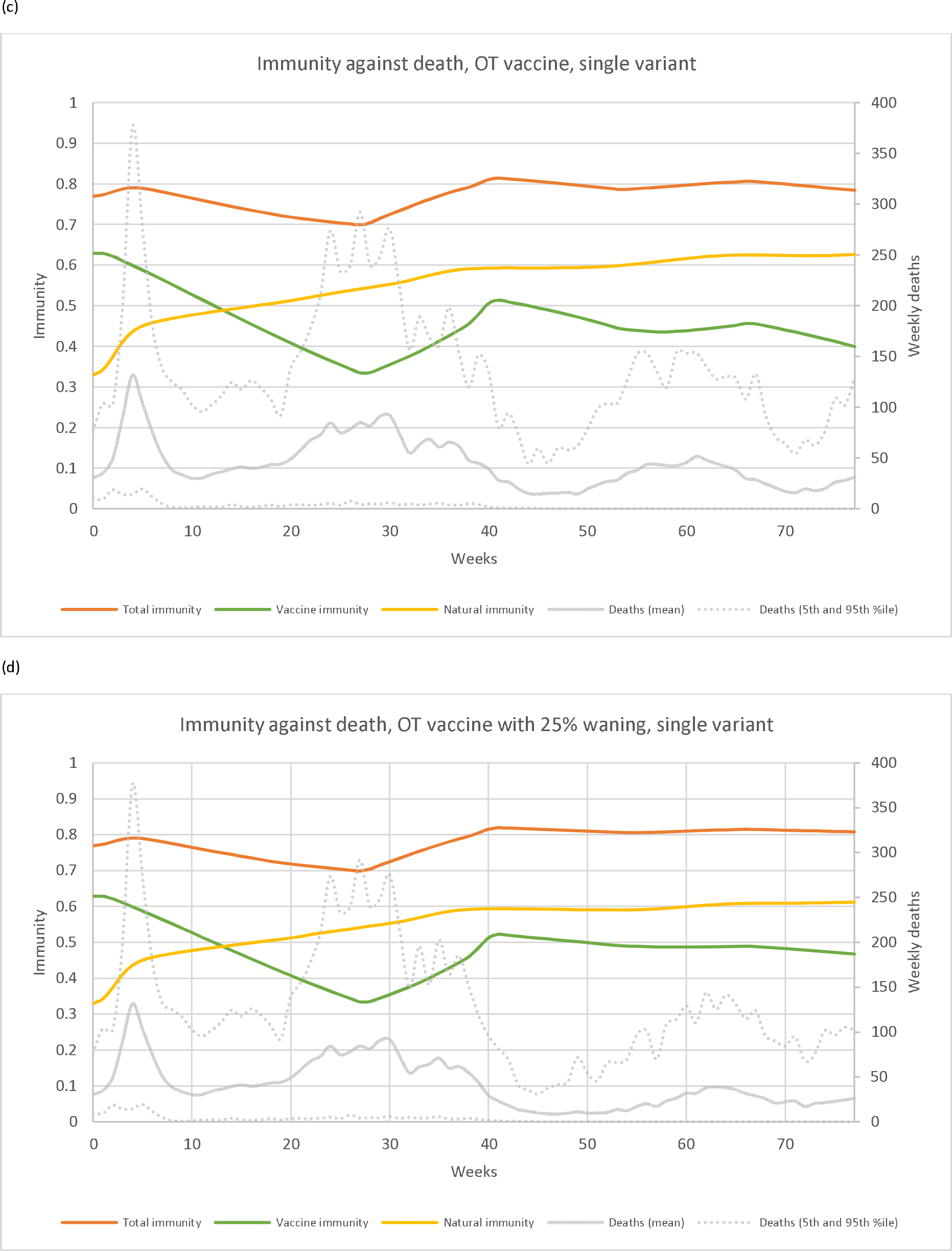
Simulated total immunity, vaccine-derived immunity, and immunity from previous infection against <u>death</u> over an 18-month period in Victoria, Australia for one future SARS-CoV-2 variant* and (a) nil further vaccination, (b) 6-monthly boosters with a current generation vaccine, (c) two doses of an Omicron-targeted next-generation vaccine with the same waning as current vaccines, and (d) two doses of an Omicron-targeted vaccine with 75% less waning than current vaccines. * variant with R_0_ 11, antigenically Omicron-like, virulence approximating the Omicron variant, emerging in July 2022 (13 weeks), with high immune escape capacity (halving vaccine effectiveness and/or natural protection on the odds ratio scale). See Table 1 for vaccination timing. CG: current generation; OT: Omicron-targeted.

SARS-CoV-2 variant (R0 11, virulence approximating that of the Omicron variant, antigenically similar to the Omicron variant, with high immune escape capacity that halves VE or natural protection on the OR scale) across the four vaccination scenarios.

With respect to total infections (Figure 2), previous infection drove most total immunity even with ongoing regular vaccination. Vaccination events provided boosts to total population immunity against infection which waned rapidly. Modelled Omicron-targeted vaccines resulted in higher peaks of total population-level immunity compared to current-generation vaccines. Waning of vaccine-derived immunity was accompanied by waves of infections followed by boosts in natural-infection derived immunity, keeping total immunity to infection relatively steady. When no further vaccines were provided, vaccine-derived immunity waned to negligible levels within months and was compensated for by natural immunity to stabilize total population-level immunity against infection. Mean cumulative infections over 18 months were 6.5 million, 6.3 million, 6 million, and 5.6 million for the nil, current generation, Omicron-targeted, and Omicron-targeted (25% waning) vaccine scenarios respectively.

For deaths (Figure 3), a greater proportion of overall immunity at the population level was accounted for by vaccination compared to that for infection (noting that waning of VE against deaths is less than that against infection). Like that seen for protection against infection, as vaccine-derived immunity to death fell, natural immunity compensated to keep overall population immunity levels relatively stable. In the scenario where no further vaccines were given beyond 1 April 2022, total immunity against death fell to 67% by 18 months; this was lower than in all scenarios of ongoing vaccination (81% at 18 months for the scenario of an Omicron-targeted vaccine with reduced waning, for example). Mean cumulative deaths over 18 months were 5,670, 3,655, 3,497, and 3,205 for the nil further, current generation, Omicron-targeted, and Omicron-targeted (25% waning) vaccine scenarios respectively.

## Discussion

Forecasting future changes in population-level immunity to SARS-CoV-2 is a complex and inherently uncertain task, made challenging by factors such as the development of new vaccines, the impact of ongoing infection, the effects of waning immunity, and the emergence of novel variants. Such analyses are critical, however, to inform decision-making. This study illustrates how population immunity is likely to vary over an 18-month period in the setting of a new SARS-CoV-2 variant and several illustrative vaccination schedules.

Our model predicts that population-level immunity will vary with SARS-CoV-2 variant characteristics. In terms of infection risk, less protection was observed at the aggregate level when the newly emergent viral variant was more virulent than current Omicron sub-variants. This reflects that for highly virulent variants hospital capacity rapidly exceeds a tolerable limit, resulting in the imposition of more stringent public health and social measures to reduce transmission. While crucial for avoiding acute morbidity and mortality and pressure on health care services from a highly virulent variant, this affords the population less protection against transmission in the future. Issues such as these highlight the complex trade-offs that characterise pandemic policy making.

For the illustrative SARS-CoV-2 variant shown in Figure 2 and Figure 3, a steady state of immunity is reached no matter the vaccination policy modelled. For protection against infection, the contribution of vaccination to overall protection in the longer-term is minimal. This differs for protection against mortality, where vaccination is responsible for the bulk of immunity early in the modelled period and provides ongoing high levels of protection in scenarios where regular vaccination continues. The marginal benefit of vaccination was observed to decrease over time, as only 80% of those who were vaccinated in the previous round receive the new vaccine (which we believe is a reasonable, if not even conservative, assumption). In Australia, 96% of eligible people received two COVID-19 vaccine doses while only 72% of those eligible have received a third dose^10^.

In situations of no further vaccination, the fall in vaccination-derived immunity is partially compensated for by natural infection immunity. This, however, is accompanied by a steadier and more sustained pattern of infections and deaths compared to the wave-like patterns for infection and mortality seen with ongoing regular vaccination. Accordingly, our modelling supports the assertion that vaccination is the preferred method of achieving maximal population immunity to SARS-CoV-2 given the risks associated with SARS-CoV-2 infection, including acute morbidity, mortality, long COVID and additional sequelae including increased cardiovascular disease risk. ^11^ Vaccination, by contrast, is associated with a very low risk of adverse effects and has the additional benefit of reducing the risk of developing long COVID. ^12^ Predictably, modelled vaccines with improved VE and less rapid waning were able to achieve similar levels of immunity to current generation vaccines but with less frequent dosing.

## Conclusions

This simulation modelling study suggests that in the absence of a new COVID-19 vaccine which provides substantially better sterilizing immunity than currently available products, the bulk of protection at the population level against SARS-CoV-2 infection is likely to continue to come from natural infection. However, vaccination appears to be responsible for a substantial amount of population-level protection against serious outcomes such as mortality. Given the risks associated with SARS-CoV-2 infection, the development of improved vaccines with enhanced protection against infection and longer lasting immunity is likely to be a critical component of the future response to the pandemic. Increasing the proportion of individuals vaccinated with each rollout would also likely assist in maintaining vaccine-mediated protection at maximum achievable levels. Finally, there is an urgent need for better estimates of protection following primary SARS-CoV-2 infection, and how these change over time and between variants. In combination with VE and waning estimates of next-generation vaccines, this will assist in informing ongoing policy decisions regarding optimal vaccination scheduling in Australia and internationally.

## Data Availability

Requests for additional model output data may be granted upon reasonable request to the researchers. Access to model code is generally not available but negotiated access may be possible. Please contact the researchers for further information.

## Acknowledgements

The authors would like to acknowledge Shania Rossiter, Samantha Howe and Jessie Zeng for their contributions during the development of the model upon which this study is based and during the conceptualization of this manuscript.

