## Supplementary material for "Relative contributions of vaccination and previous infection to population-level SARS-CoV-2 immunity over time: a simulation modelling study"

*Supplementary Table 1: Public health and social measures by stage (1 to 5)*

| | Stage 1 | Stage 2 | Stage 3 | Stage 4 | Stage 5 |
|---|---|---|---|---|---|
| Proportion of people who try to avoid contact with others (excluding their household)†* | 35% | 53% | 63% | 75% | 85% |
| Proportion of time spent trying to avoid contacts, for those that attempt to do so†* | 45% | 63% | 73% | 83% | 90% |
| Proportion of workers attending work in person† | 70% | 51% | 35% | 20% | 10% |
| Schools open (disable contact avoiding behaviour among students) | Yes | Yes | Yes | No | No |
| Proportion of people that wear masks outside the home† | | | | | |
| ≥20-year-olds | 20% | 35% | 60% | 80% | 90% |
| 10- to 19-year-olds | 16.7% | 26.67% | 40% | 53.3% | 60% |
| <10-year-olds | 11.1% | 17.78% | 26.67% | 35.6% | 40% |
| Proportion of mask wearing that is with a respirator# | 20% | 20% | 20% | 20% | 20% |
| Proportion of people that engage in super spreading behaviour each day (move to a random gathering location)† | 5% | 3.5% | 2.2% | 1.6% | 1% |
| Underlying frequency of visiting a random nearby gather location each day (e.g., a supermarket) | 14.28% | 14.28% | 13% | 7.5% | 5% |
| Radius for determining whether a gather location counts as nearby | 8.8 | 6.7 | 6 | 5 | 3.6 |
| Maximum distance moved by an agent each day | 12 | 9 | 7.5 | 6 | 5 |

*†ORs of 2, 4, 6 and 8 are applied to the proportion of people that wear a mask, isolation compliance, proportion of people that avoid others, and proportion of time spent avoiding others for those aged 50-60, 60-70, 70-80 and ≥80 respectively to capture increasing infection avoidance behaviour in older age groups. Reciprocals of these ORs are applied to the daily chance of visiting a gather location, and daily chance of superspreading. Note agents ≥60 years old are excluded from the category of workers.*

**In stages 1-3, the proportion of people who try to avoid contact with others and the proportion of time spent trying to avoid contacts for those attempting to do so are dynamic, based on the average number of infections over the last 7 days. Proportions are as written above if the 7-day average of infections is <5000. Proportion of people who try to avoid contacts and proportion of time spent avoiding increase to a maximum of 15 and 10 percentage points higher than those written above, respectively, at 32,000 daily infections over the last seven days. The region between 5000 and 32,000 is linearly interpolated.*

*#Only applies to those aged ≥10 years.*

*Supplementary Table 2: (De)escalation rules for stages of public health and social measures, based on projected hospital capacity*

**Triggers**

Escalation:

If average expected number of people in hospital due to COVID-19 10-14 days (inclusive) into the future is:

- >300 per million → Stage 5
- >200 per million → Stage 4
- >130 per million → Stage 3
- >90 per million → Stage 2

De-escalation:

If no de-escalation in last 7 days, and average expected number of people in hospital due to COVID-19 10-14 days (inclusive) into the future is:

- <230 per million → Stage 4 if Stage 5
- <150 per million → Stage 3 if in stage 4 or 5
- <100 per million → Stage 2 if in Stage 3, 4 or 5
- <70 per million → Stage 1

*Supplementary Table 3: Immune escape odds ratios, vaccine effectiveness*

| | **Current generation vaccine, 2 doses** | **Current generation vaccine, ≥3 doses** | **Omicron-targeted vaccine, ≥1 dose** |
|---|---|---|---|
| Omicron BA.2 | 1 (ref)† | 1 (ref)‡ | *2* |
| Omicron-like variant with $R_0$ 11 | *0.707, 0.5* | *0.707, 0.5* | *1.414, 1* |
| Omicron-like variant with $R_0$ 14 | *1, 0.707* | *1, 0.707* | *2, 1.414* |

*†Reference for the current generation double dose column.*

*‡Reference for the current generation triple dose and next-generation omicron-targeted columns.*

*These odds ratios (ORs) apply at any point in time, with waning post last dose also in effect. All italicized ORs are drawn from a range of 0.841 to 1.189 of their stated values, with 100% correlation (including the OR of 2 for next-generation vaccines against current Omicron) uniformly on the log OR scale. For each combination of vaccine and variant, 2 possible levels of possible immune escape are modelled. For example, for the variant referred to in Figures 2 and 3, the lower (i.e., higher immune escape) of the 2 values in each cell for an Omicron-like variant with an $R_0$ of 11 apply.*

*Supplementary Table 4: Immune escape odds ratios, infection-derived immunity*

| | **Previous infection with Omicron BA.2** | **Previous infection with new variant** |
|---|---|---|
| Omicron BA.2 | 1 (ref) | *1, 1* |
| Omicron-like variant with $R_0$ 11 | *0.707, 0.5* | *1, 1* |
| Omicron-like variant with $R_0$ 14 | *1, 0.707* | *1, 1* |

*These odds ratios (ORs) apply at any point in time, with waning post previous infection also in effect. All italicized ORs are drawn uniformly from a range of 0.841 to 1.189 of their stated values, with 100% correlation, uniformly on the log OR scale. For each combination of previous/current variant, 2 levels of possible immune escape are modelled.*

*Supplementary Figure 1: Vaccine effectiveness estimates for triple dose of current generation vaccine by clinical outcome, individuals aged <60 years, Omicron BA.2 variant.*

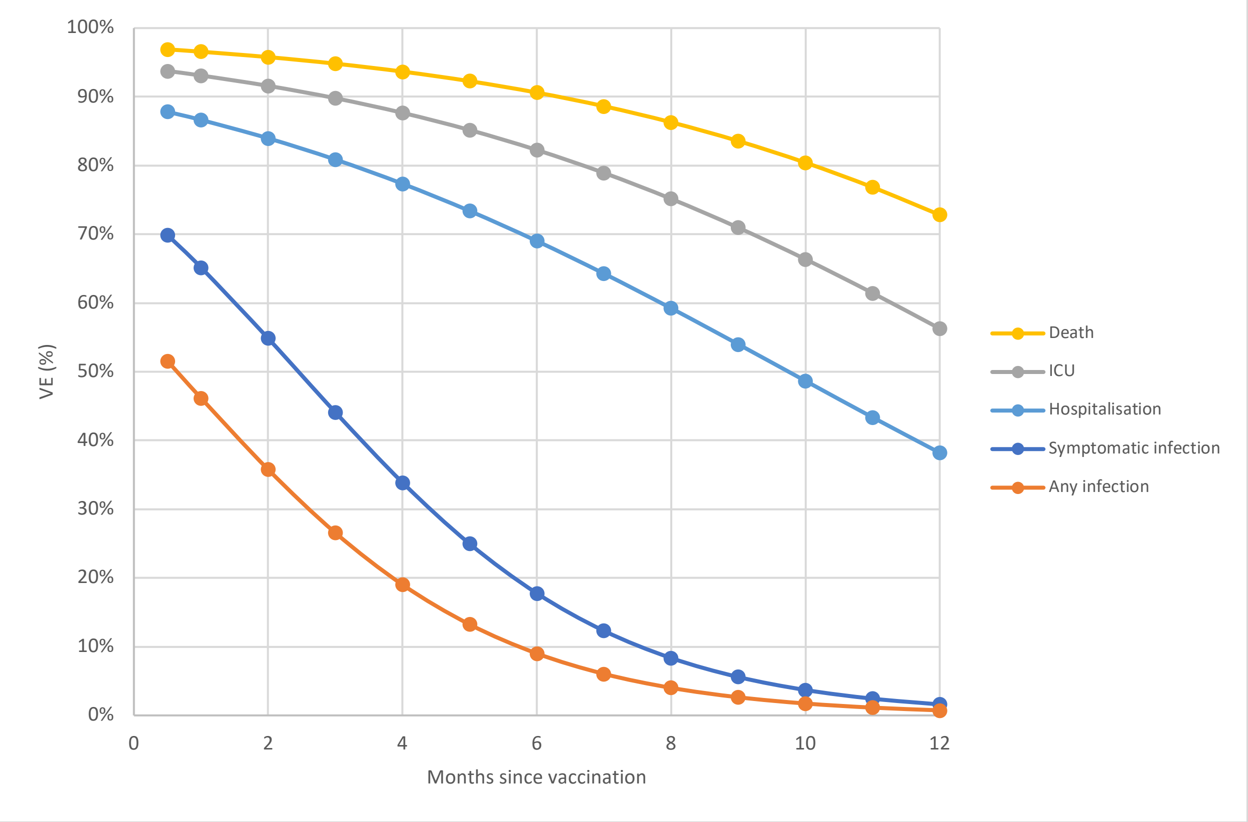


*Odds ratios for effectiveness of Omicron-targeted vaccines and immune escape apply to these estimates to calculate the final level of protection. Vaccine effectiveness estimates vary by age. A similar system (with different levels of protection) applies for immunity following infection. For details see the supplementary materials of* [1]
